# Faecal shedding models for SARS-CoV-2 RNA among hospitalised patients and implications for wastewater-based epidemiology

**DOI:** 10.1101/2021.03.16.21253603

**Authors:** Till Hoffmann, Justin Alsing

## Abstract

**Summary:** The concentration of SARS-CoV-2 RNA in faeces is not well established, posing challenges for wastewater-based surveillance of COVID-19 and risk assessments of environmental transmission. We develop versatile hierarchical models for faecal RNA shedding and apply them to data collected in six studies. We find that the mean number of gene copies per mL of faeces is 1.9 × 10^6^ (2.3 × 10^5^–2.0 × 10^8^ 95% credible interval) among unvaccinated hospitalised patients. Using Bayesian model comparison, we find no evidence for a subpopulation of patients who do not shed RNA: limits of quantification can account for negative stool samples. Our models indicate that hospitalised patients represent the tail of the shedding profile with a half-life of 34 hours (28–43 95% credible interval), suggesting that wastewater-based surveillance signals are more indicative of incidence than prevalence and can be a leading indicator of clinical presentation. Shedding among inpatients cannot explain high RNA concentrations observed in wastewater, consistent with more abundant shedding during the early infection course. We show that the models generalise and can predict summary statistics of held-out clinical datasets. However, shedding prior to hospitalisation cannot be constrained due to lack of samples, and information on viral variants was not available.

## 1. Introduction

The novel virus SARS-CoV-2 (severe acute respiratory syndrome coronavirus 2) has infected over 500 million people, and more than six million people have succumbed to the resultant disease, COVID-19 (coronavirus disease 2019) (Dong et al., 2020). Individuals infected by the virus primarily suffer from respiratory symptoms (Huang et al., 2020), but gastrointestinal manifestations of the disease have also been observed (Cheung et al., 2020). The presence of viral RNA in faeces allows for the surveillance of COVID-19 by quantifying gene copies in sewage (Medema et al., 2020). So-called wastewater-based epidemiology (WBE) provides data that can complement traditional testing schemes and can be used to monitor the disease relatively cheaply by pooling wastewater from thousands of people (Polo et al., 2020). Correlations between case numbers from individual testing schemes and RNA concentrations in wastewater have been observed (Medema et al., 2020; Wu et al., 2022; Morvan et al., 2022). However, associative studies cannot easily be used to calibrate WBE approaches: each sewerage system is different (Banks et al., 2018), and case numbers may not be a good indicator of prevalence (Slot et al., 2020)—especially when testing capacity is limited. The World Health Organisation considers “quantitative information on viral shedding” an imminent need to reap the potential benefits of WBE (Global Infectious Hazard Preparedness Team, 2020, p. 3).

Faecal shedding of RNA suggests that the virus could be transmissible via the faecal-oral route (Wang et al., 2020). While presence of RNA does not imply presence of infective virus, the likelihood increases with higher RNA loads (Wölfel et al., 2020). Sewer overflows could cause spillover events, leading to new viral reservoirs (Franklin and Bevins, 2020). For example, mink are susceptible to SARS-CoV-2 (Oreshkova et al., 2020) and can be exposed to untreated sewage (Franklin and Bevins, 2020). Furthermore, wastewater workers are at risk of contracting sewage-borne pathogens (Zabinski et al., 2018), and wastewater is a possible infection mode in densely populated communities (Kang et al., 2020). Quantifying these risks is essential for making informed policy decisions.

We developed a family of random-effect models (Gelman et al., 2014a, ch. 5) for SARS-CoV-2 RNA concentrations in faecal samples and applied them to data from six clinical studies to study three aspects of faecal RNA shedding. First, we studied the shedding profile, i.e. the temporal variability of shedding over the infection course, which affects the interpretation of WBE results (Wu et al., 2022). We find that the profile decays quickly with a half-life of 34 hours. Second, the proportion of patients with one or more positive faecal samples has been extensively studied (Cheung et al., 2020; van Doorn et al., 2020; Wong et al., 2020). We determined that the limit of quantification of assays can account for patients without positive samples. Bayesian model comparison revealed no evidence for a subpopulation of patients who do not shed RNA faecally. Third, we obtained estimates of the mean faecal RNA concentration: a quantity important for inferring disease prevalence or incidence from wastewater data (Ahmed et al., 2020). We show that the models are able to predict summary statistics of held-out studies accurately and consider the implications of our results for wastewater-based epidemiology.

## 2. Methods

### 2.1. Data acquisition

We searched the literature for primary research that reported quantitative information on SARS-CoV-2 RNA loads in faeces, and we identified eleven studies of interest (Cheung et al., 2020; Wang et al., 2020; Wölfel et al., 2020; Han et al., 2020; Kim et al., 2020a; Ng et al., 2020; Zheng et al., 2020; Jeong et al., 2020; Zhang et al., 2020; Pan et al., 2020; Lui et al., 2020). Semi-quantitative studies reporting only cycle-threshold values were excluded.

Data from four studies were used to fit the hierarchical models described in section 2.2. First, Wölfel et al. (2020) reported longitudinal faecal RNA concentrations for nine patients in Munich who had been in close contact with a common index case. Second, Lui et al. (2020) followed the first eleven patients hospitalised due to COVID-19 in Hong Kong. Third, Han et al. (2020) studied the viral dynamics of a neonate and her mother in Seoul. Fourth, Wang et al. (2020) reported cycle threshold (*C*_*t*_) values of RTqPCR assays for faecal samples from a subset of patients admitted to three hospitals in China, and we transformed *C*_*t*_ values to RNA concentrations using conversion constants provided in the publication. Longitudinal information was not available. Table 1 lists information on the number of patients and samples for each study.

**Table 1.** Six studies providing quantitative data on faecal RNA loads were analysed. Microdata, i.e. sample-level RNA concentrations, provided by the first four studies were used to fit random effects models. The next two studies did not provide microdata, and we validated our models by comparing reported summary statistics with model-based predictions, as discussed in section 2.4. Samples with viral loads below the limit of quantification (LOQ) are considered negative for SARS-CoV-2. The table also reports the total number of patients *n* and number of samples *m*, including a breakdown by positivity.

| study | patients | | | samples | | | micro-<br>data | longi-<br>tudinal | LOQ<br>($\log_{10} \text{ mL}^{-1}$) |
| --- | --- | --- | --- | --- | --- | --- | --- | --- | --- |
| | $n$ | + | − | $m$ | + | − | | | |
| Wang et al. (2020) | 14 | 6 | 8 | 14 | 6 | 8 | yes | no | 1.4 |
| Wölfel et al. (2020) | 9 | 8 | 1 | 82 | 68 | 14 | yes | yes | 2.0 |
| Lui et al. (2020) | 11 | 11 | 0 | 43 | 23 | 20 | yes | yes | 2.8 |
| Han et al. (2020) | 2 | 2 | 0 | 9 | 8 | 1 | yes | yes | 3.8 |
| Kim et al. (2020a) | 38 | 8 | 30 | 129 | 13 | 116 | no | no | ? |
| Ng et al. (2020) | 21 | 21 | 0 | 81 | ? | ? | no | no | 2.5 |

Two studies provided summary statistics of viral RNA concentrations in faecal samples, such as the highest concentration observed, together with sufficiently detailed descriptions of the study design to replicate the studies *in silico*, as described in section 2.4.

In particular, Kim et al. (2020a) quantified RNA concentrations in faecal samples collected from hospitalised patients, and Ng et al. (2020) studied RNA concentrations in faecal samples to assess transmission risks associated with faecal microbiota transplants. These data are not used to fit the hierarchical models but instead to assess the out-of-sample predictive utility of our fitted models.

Five studies were excluded from the analysis despite providing quantitative information. First, using an assay originally developed for samples from ferrets (Kim et al., 2020b), Jeong et al. (2020) reported RNA concentrations of various specimen types, including faecal samples. However, the reported concentrations are orders of magnitude lower than reported in any of the other quantitative studies for any specimen type. Indeed, most measurements are below the limit of quantification of other studies. Second, Zhang et al. (2020) reported detailed data in a figure, but it is not possible to extract the relevant information because of the low resolution of the figure, and the data could not be obtained by other means. Third, Zheng et al. (2020) conducted a large study, comprising 85 patients and 842 faecal samples. Unfortunately, viral loads are only reported gas the median for each patient (private communication). Finally, Pan et al. (2020) and Cheung et al. (2020) collected faecal RNA concentration data from 17 and 59 confirmed cases, respectively, but neither study provides information on the number of samples collected.

### 2.2. Models

To study faecal RNA shedding quantitatively, we developed a suite of hierarchical models for RNA concentrations in faecal samples. In contrast to existing quantitative approaches (Miura et al., 2021; Benefield et al., 2020), our models can account for a variable number of samples per patient, incorporate data from studies with different levels of quantification, and capture variability between patients as well as variability between samples from the same patient. The baseline model assumes that all infected patients shed viral RNA faecally and that typical RNA concentrations in samples vary over the course of the infection. We considered three different shedding profiles: an exponential decay profile, a gamma profile, and the exponential rise-and-decay profile proposed by Teunis et al. (2015) for norovirus RNA shedding (see section 2.2.1 for details). We call this the *temporal standard* model, and it is illustrated in fig. 1.

**Fig. 1.**
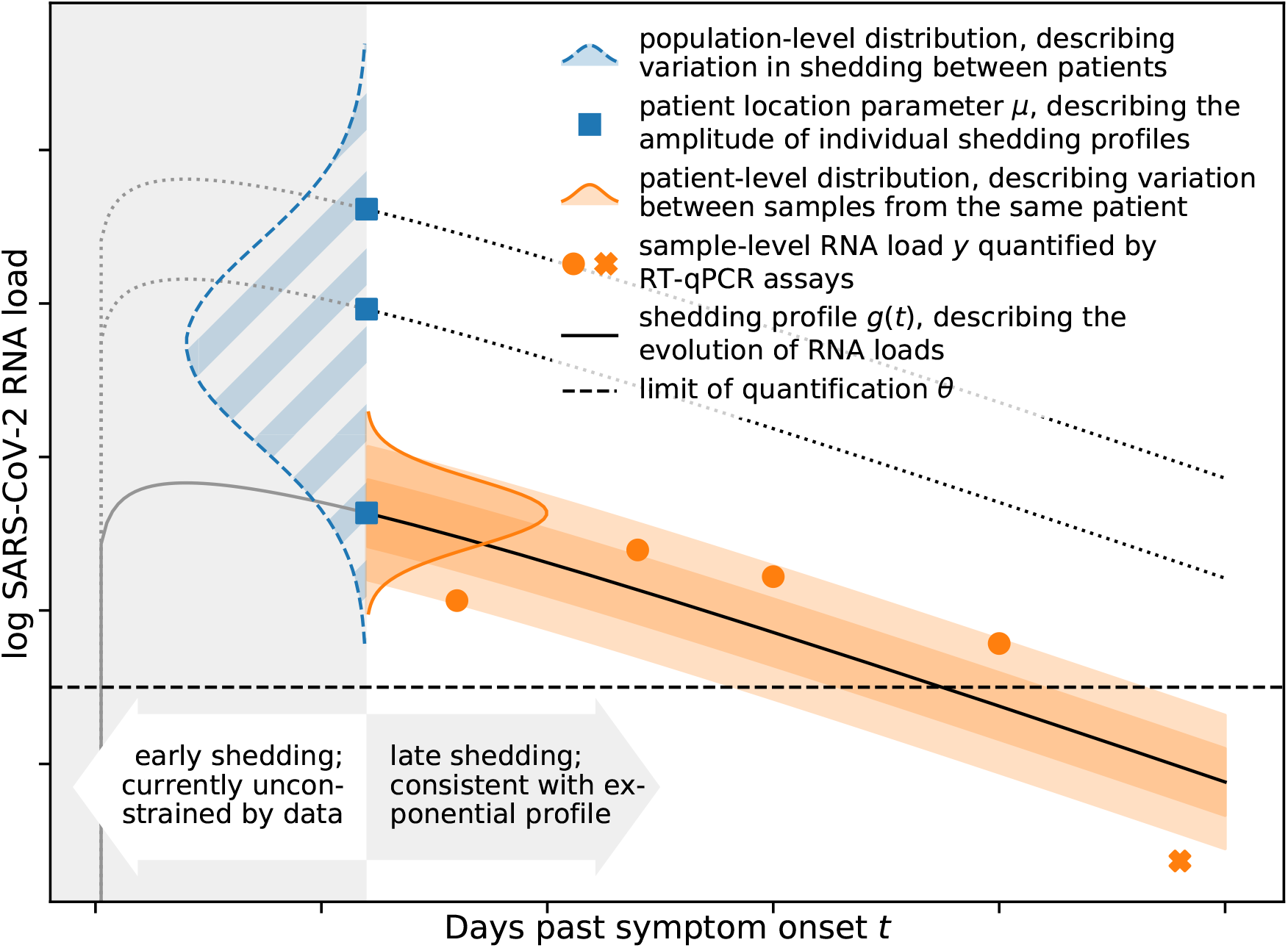
A flexible hierarchical model can capture a wide range of aspects of real-world data. The population-level distribution (hatched) captures variation in shedding behaviour between patients, giving rise to location parameters *μ*_*i*_ for each patient *i* (see section 2.2 for details). The location parameters (squares) describe the amplitude of individual shedding profiles as illustrated for three patients. The shedding profile (solid line) modulates typical RNA concentrations in faecal samples over time, and the patient-level distribution (shown for one patient) describes the variation between samples from the same patient. All samples are analysed using an RTqPCR assay with a given limit of quantification (LOQ) *θ* (dashed line), and concentrations above the LOQ (circles) can be quantified. Concentrations below the LOQ (cross) cannot be quantified. Currently available data do not allow us to constrain the early shedding profile, and late-stage shedding is consistent with an exponential decay profile.

We considered two modifications to the baseline model. First, we considered a *constant* model, where the time-variability of shedding was removed. Second, to assess whether there exists a subpopulation of patients who never shed viral RNA faecally,

we introduced a shedding prevalence parameter *ρ* such that all samples of an infected patient are negative with probability 1 − *ρ* (see section 2.2.2 for details). We call this the *subpopulation* model (as opposed to the *standard* model in which all patients shed RNA faecally). Combining the two modifications (with and without a subpopulation of non-shedders, and with and without time-variability) gives rise to four models in total.

We begin by describing the simple *constant standard* model in three steps because of its relative simplicity. First, the mean faecal RNA concentration *λ*_*i*_ for each patient *i* ∈ {1, …, *n*} follows a distribution with probability density function *f*, where *n* is the number of patients. Log-normal, Weibull, and gamma distributions are common choices for distributions that model positive, continuous data (Kappenman, 1985). However, conclusions based on these distributions can differ substantially due to their tail behaviour (Rubin, 1984). We thus employed a generalised gamma distribution (GGD) with shape *Q*, location *M*, and scale *S*. The parameters *M* and *S* can be understood as location and variability on the natural log scale, respectively. The GGD is a flexible distribution which encompasses the log-normal distribution (*Q* = 0), Weibull distribution (*Q* = 1), and gamma distribution (*Q* = *S*) as special cases (Prentice, 1974). This choice comes at the cost of wider credible intervals, commensurate with our lack of prior knowledge about the shape of the shedding distribution.

Second, *m*_*i*_ samples are collected from each patient *i*. The RNA concentration *y*_*ij*_ in sample *j* from patient *i* follows a GGD with shape *q*, location *μ*_*i*_, and scale *σ*. For the *constant* model (without time-varying shedding), the location parameter *μ* for the patient-level distribution is chosen such that ⟨*y*_*ij*_⟩= *λ*_*i*_ for each patient. Because of the properties of the generalised gamma distribution, we can express the location parameter in terms of the mean as

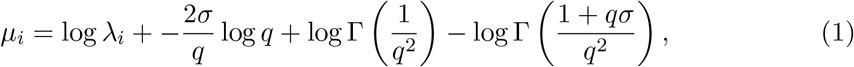

where Γ denotes the gamma function (see the Appendix for details).

Third, the RNA concentration *y*_*ij*_ is quantified using an assay with level of quantification (LOQ) *θ*_*ij*_ (see Kitajima et al. (2020) for an overview of different assays). If the concentration is below the LOQ, the sample is considered negative for the purpose of this study, and we denote the output of the assay as *x*_*ij*_ = °. Otherwise, the result of the assay faithfully captures the RNA concentration in the sample, i.e. *x*_*ij*_ = *y*_*ij*_. We do not explicitly model measurement error or variability between technical replicates in this study because the relevant data are not available and assays tend to yield reproducible results. For example, the CDC N1 assay (Lu et al., 2020) has a coefficient of variation *<* 3.7%—much smaller than typical variability between samples from the same patient. The RNA quantification is censored by the LOQ, and the likelihood of observing a particular assay result *x*_*ij*_ is thus

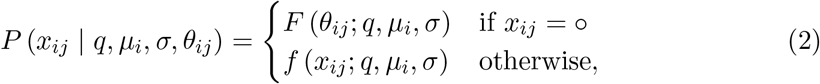

where *f* denotes the probability density function of the generalised gamma distribution and *F* denotes the corresponding cumulative distribution function (Prentice, 1974).

#### 2.2.1. Time-variation

RNA shedding varies over the course of the infection (Cevik et al., 2020), and the *constant* model cannot capture these changes. We incorporate temporal variability by introducing a shedding profile such that the expected RNA concentration *λ*_*i*_ for each patient *i* varies over time. In particular, we let *λ*_*i*_(*t*) = *λ*_*i*_(*t* = 0)*g*(*t*), where *t* is the number of days since symptom onset, *g*(*t*) is the shedding profile that modulates the expected RNA concentration, and *λ*_*i*_(*t* = 0) is sampled from the population-level distribution. The shape and scale parameters *q* and *σ* of the patient-level distribution are kept constant. Because all available data were collected from hospitalised patients several days after the initial onset of symptoms, we can only constrain the later part of the shedding profile. We used an exponential profile *g*_exp_(*t*) = exp (−*αt*), where *α* is the decay rate of the profile, because it provides an adequate fit for late-stage faecal shedding of other viruses (Teunis et al., 2015). Substituting into eq. (1), the exponential shedding profile gives rise to a location parameter that varies linearly with the number of days after symptom onset for each patient *i* such that *μ*_*i*_(*t*) = *μ*_*i*_(*t* = 0) − *αt*.

Any reporting error associated with the number of days since symptom onset *t* can be compensated for by a corresponding change in *μ*_*i*_(*t* = 0). This explains why the inferred shedding profile is robust to reporting inaccuracies, as discussed in section 3. The profile shown in fig. 2 (b) is exp (*M* − *αt*), and it captures the decay of the effective population-level location parameter *M* over time. The profile half life is *τ*_1*/*2_ = log(2)*/α*. We also considered two further shedding profiles to assess the sensitivity of our inferences to the choice of *g*(*t*). First, we used a gamma shedding profile

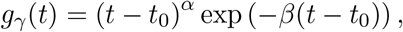

where *α >* 0 and *β >* 0 control the shape of the profile and *t*_0_ is the time at which shedding can first occur, i.e. *g*(*t < t*_0_) = 0. Second, we considered the exponential rise-and-decay profile proposed by Teunis et al. (2015) for norovirus shedding

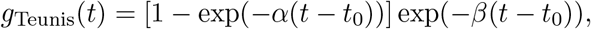

where *α* and *β* control the rise and decay of the profile, respectively.

**Fig. 2.**
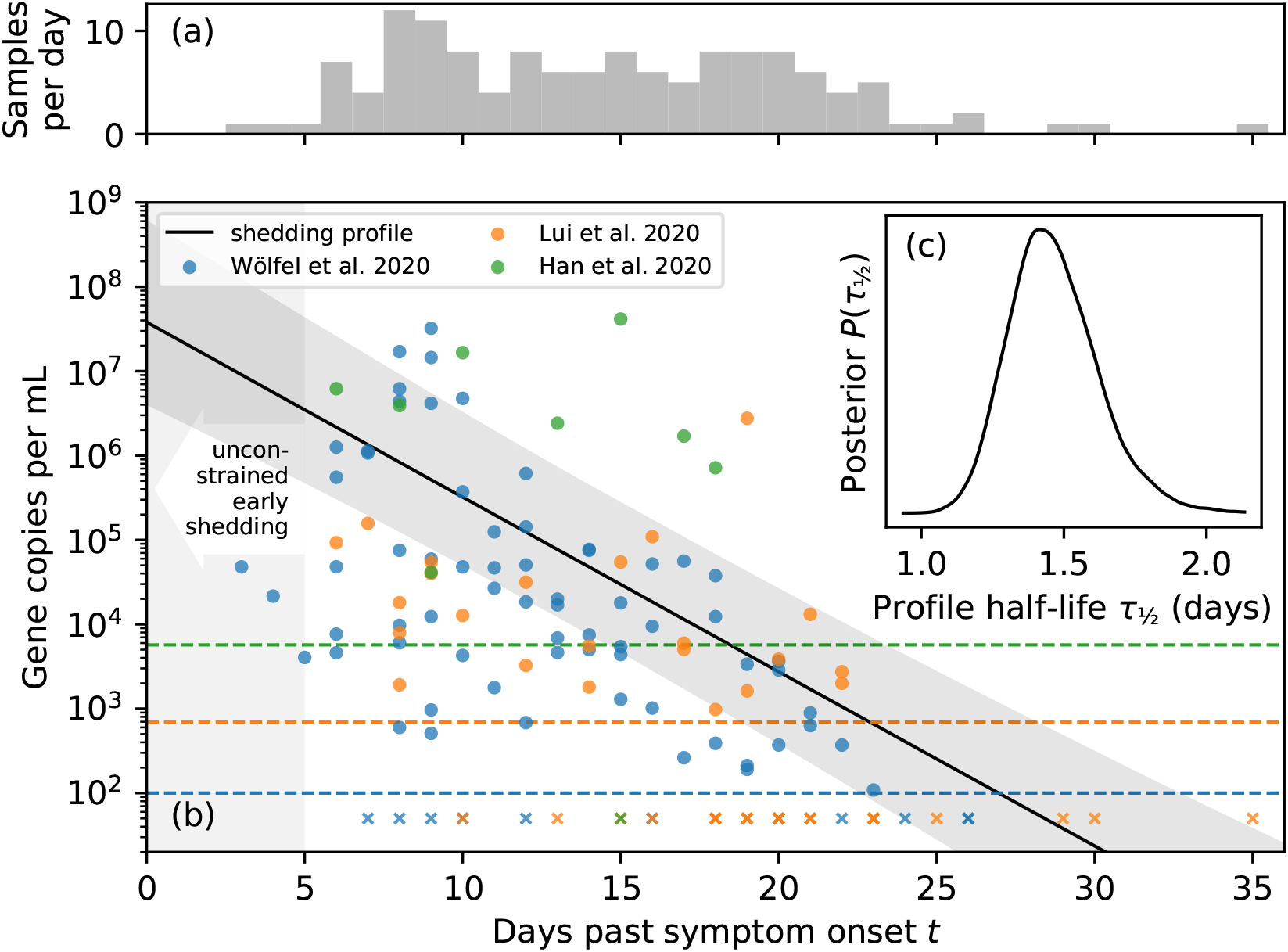
Faecal RNA concentrations decay rapidly over the infection course. Panel (a) shows a histogram of the number of samples collected on each day post symptom onset. The small number of samples collected within the first six days after symptom onset and the absence of samples collected prior to symptom onset make it difficult to constrain shedding during the early infection course. Panel (b) shows longitudinal faecal RNA concentration data from three studies together with the level of quantification (LOQ) for each study as dashed lines. Circles and crosses represent samples above and below the LOQ, respectively. The time-dependent exponential shedding profile of the *temporal standard* model is shown in black, and the shaded region represents the 95% credible interval of the profile (see section 2.2.1 for details). Panel (c) shows the posterior distribution for the half-life *τ*_1*/*2_ of the shedding profile.

#### 2.2.2. Non-shedding subpopulation

To investigate whether there is a subpopulation of patients who do not shed SARS-CoV2 RNA faecally, we extended the model by introducing a binary indicator *z*_*i*_ ∈ {0, 1} for each patient *i*. If *z*_*i*_ = 1, the shedding behaviour of the patient is unchanged. If *z*_*i*_ = 0, patient *i* does not shed RNA, and *x*_*ij*_ = for all samples *j*. The indicator variables follow a Bernoulli distribution with probability *ρ*, i.e. the prevalence of shedding among the population of patients. This extension gives rise to what we refer to as the *subpopulation* models.

We know that *z*_*i*_ = 1 for any patient with one or more positive samples, and the likelihood follows eq. (2). However, for any patient *i* whose *m*_*i*_ samples 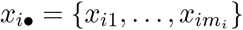 are all below the LOQ, the likelihood is

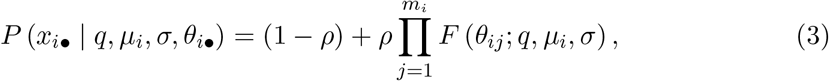

where the first term accounts for a patient who cannot shed RNA (*z*_*i*_ = 0) and the second accounts for a patient whose samples are all below the LOQ (*z*_*i*_ = 1).

### 2.3 Parameter inference and model comparison

We used the nested sampler *polychord* (Handley et al., 2015) which draws samples from the posterior distribution and evaluates the model evidence, i.e. the marginal likelihood of the data under the model, as listed in table 2. The study by Wang et al. (2020) was not used to estimate parameters for *temporal* models because longitudinal data were not available. Similarly, to facilitate a direct comparison between models, that study was also omitted from the evaluation of evidences. We used unit-scale half-Cauchy prior distributions for the shape parameters *Q* and *q* as well as the scale parameters *S* and *σ*. A flat prior on the interval 6–23 was used for the population location *M*. This corresponds to a flat prior on 2.6–10 on the log_10_ scale which is sufficiently wide to avoid boundary effects. A flat prior on the unit interval was used for the shedding prevalence *ρ*. For models with temporal variability, a unit-scale Cauchy prior was used for the profile parameters *α* and *β*. We used uniform prior on the interval -14–7 for *t*_0_ such that the onset of shedding can differ from the onset of symptoms. All inferences were run in triplicate, and no sensitivity to different pseudo-random number generator seeds was observed (Gelman-Rubin diagnostic 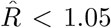 for all models (Gelman and Rubin, 1992)).

**Table 2.** Accounting for temporal variability of the shedding profile is essential, and there is no evidence for a subpopulation of patients who do not shed RNA faecally. Model evidences evaluated on three common datasets prefer *temporal* models (evidence shown for an exponential decay profile) over *constant* ones, and *standard* models are preferred over models with a *subpopulation* of patients who do not shed SARS-CoV-2 RNA faecally (higher is better). Parameter estimates are consistent with conclusions based on model evidences. They are reported as the posterior mode together with the 95% credible interval in brackets. All reported credible intervals are highest posterior density intervals. Parameter estimates for *constant* models include a fourth dataset without temporal information. The adjusted Watanabe-Akaike information criterion (WIAC) strongly prefers *temporal* models and marginally favours models with a *subpopulation* of patients who do not shed (lower is better). Errors for evidences and WAIC are standard errors, and “zero” errors indicate errors less than the reported number of digits.

| tem-<br>poral | sub-<br>pop. | log evidence | mean conc.<br>($\log_{10} \text{ mL}^{-1}$) | half-life $\tau$<br>(hours) | shedding<br>prevalence $\rho$ | adjusted<br>WAIC |
| --- | --- | --- | --- | --- | --- | --- |
| no | no | $-1303.1 \pm 0.2$ | 6.28 [5.36, 8.30] | constant | 1 (by defn.) | $390.5 \pm 0.0$ |
| no | yes | $-1305.5 \pm 0.2$ | 6.28 [5.40, 8.04] | constant | 0.95 [0.74, 1.00] | $390.0 \pm 0.0$ |
| yes | no | $-1269.3 \pm 0.2$ | variable | 34 [28, 43] | 1 (by defn.) | $386.7 \pm 0.1$ |
| yes | yes | $-1271.6 \pm 0.2$ | variable | 34 [28, 43] | 0.97 [0.82, 1.00] | $386.0 \pm 0.1$ |

In addition to the results discussed in section 3, we consider four technical implications of the inference here. First, the gamma and Teunis shedding profiles are both consistent with the exponential shedding profile for late-stage shedding where data are available to constrain them. However, the available data cannot constrain early shedding behaviour, as illustrated by the wide range of shedding profiles consistent with the data shown in fig. 3. Miura et al. (2021) reported tight posteriors for early shedding behaviour using the shedding profile proposed by Teunis et al. (2015), but their inferences rely on the assumption that the time of symptom onset and shedding onset coincide (i.e. *t*_0_ = 0), an assumption that is not yet supported by evidence.

**Fig. 3.**
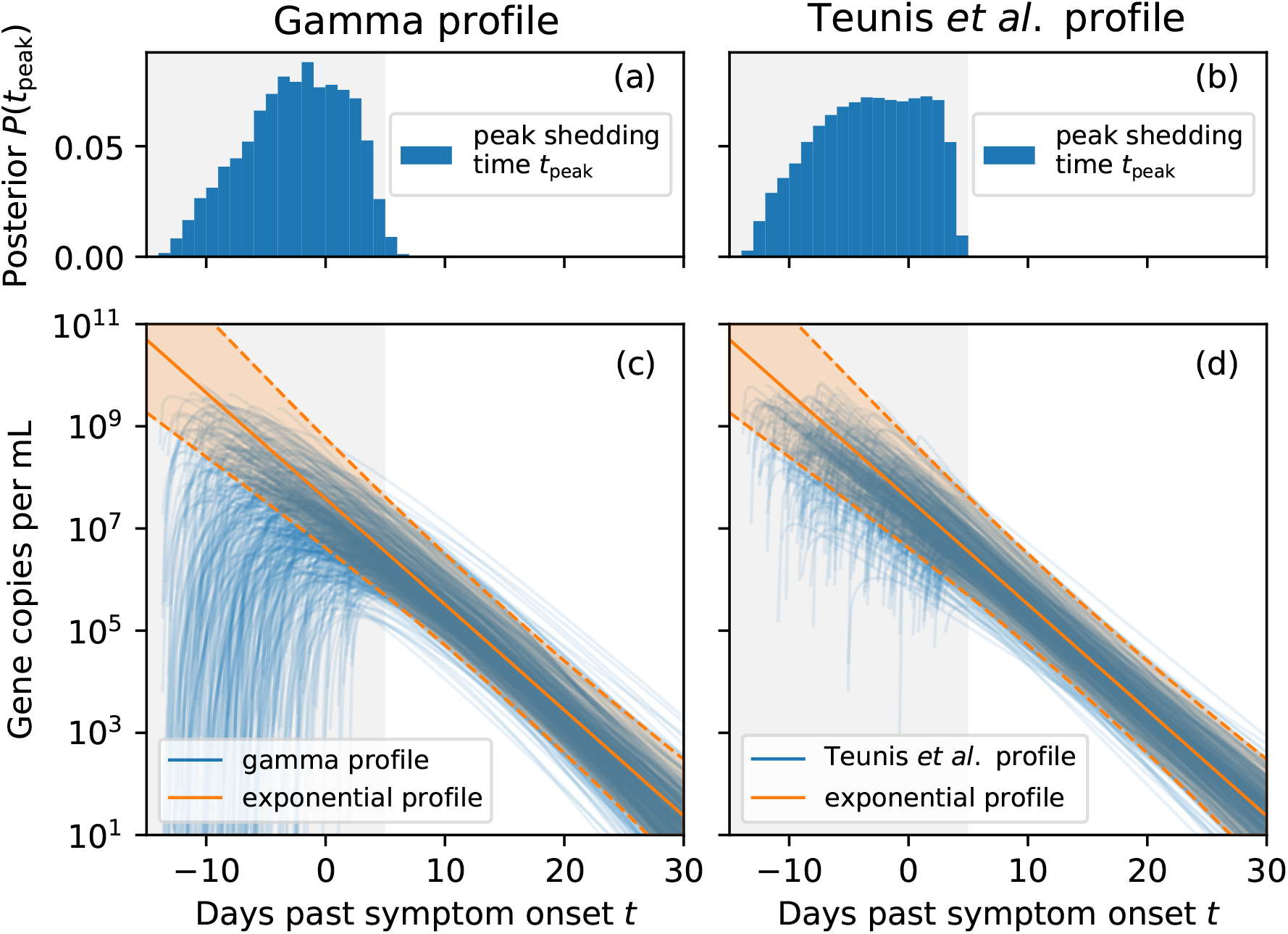
Different shedding profiles yield consistent results for late-stage shedding. But early shedding behaviour (prior to day five after symptom onset) cannot be constrained given the available data, including peak RNA concentration. Panels (a) and (b) show the the posterior distributions for the time *t*_peak_ at which the inferred gamma and Teunis shedding profiles peak, respectively. Because the data cannot constrain early shedding, the posterior for *t*_peak_ has broad support. Panels (c) and (d) show posterior samples of the gamma and Teunis profiles, respectively, as individual lines. The exponential profile discussed in section 2.2.1 is overlaid as a solid line, and the shaded region corresponds to the 95% credible interval.

Second, the shape *Q* controls the tails of the population-level distribution: the larger *Q*, the lighter the tails (see Prentice (1974) for details). The population-level mean RNA concentration, i.e. the expected RNA concentration in a random faecal sample from a previously unobserved patient, depends on the tail behaviour because the mean is sensitive to outliers (Rubin, 1984). As shown in fig. 4 (a), the inferred mean under the *constant standard* model is larger for smaller *Q* because of the heavier tails. The corresponding credible intervals are also wider because constraining the mean requires more data for heavier-tailed distributions. Employing log-normal, Weibull, or gamma distributions would have entailed a poorly-motivated implicit choice about the shape of the distribution. Using generalised gamma distributions instead allows us to explicitly account for our prior uncertainty about the shape of the tails.

**Fig. 4.**
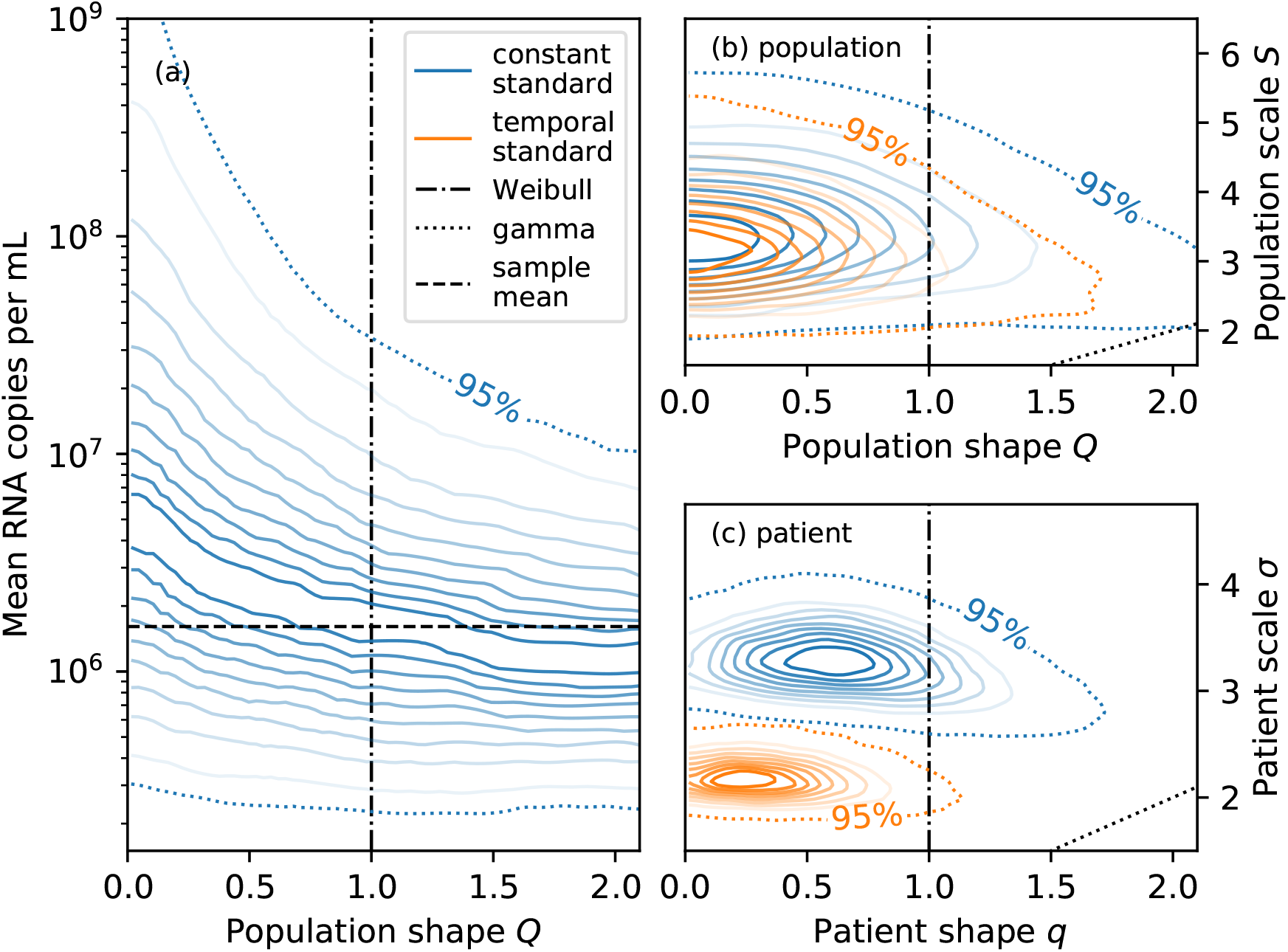
The mean number of RNA copies per mL of faeces is sensitive to the shape of the shedding distribution. Panel (a) shows the conditional distribution of the mean number of RNA copies per mL of faeces given the population shape parameter *Q* for the *constant standard* model. The mean tends to be larger for smaller *Q* due to heavier tails. Panels (b) and (c) show the joint distributions of the shape and scale parameters for the population-level distribution and patient-level distribution, respectively. Parametrisations corresponding to the Weibull distribution (*Q* = *q* = 1) and gamma distribution (*Q* = *S* and *q* = *σ*) are shown as black dot-dashed and dotted lines, respectively. The patient-level scale parameter *σ* is larger for the *constant* than the *temporal* model (both without a non-shedding subpopulation) because the patient-level distribution needs to account for both temporal variability and idiosyncratic sample-to-sample variability.

Third, the 95% credible region for the population-level shape *Q* and scale *S* is consistent with the log-normal distribution (*Q* = 0) or Weibull distribution (*Q* = 1) for both the *constant* and *temporal* models, as shown in fig. 4 (b). However, the special case of the gamma distribution (*Q* = *S*) can be confidently excluded because its tails are too light. The same conclusions apply to the patient-level distribution.

Fourth, the joint posterior distribution for the population-level shape and scale parameters are similar under both the *constant* and *temporal* models (exponential shedding profile) without a subpopulation of non-shedders, as shown in fig. 4 (b): modelling the temporal shedding profile does not have a significant effect on the inferred variability in shedding behaviour between patients. However, as shown in fig. 4 (c), the patient-level scale *σ* is significantly larger for the *constant* than the *temporal* model. The *constant* model can only account for large (time-integrated) sample-to-sample variability with a broad distribution. In contrast, the *temporal* model can explain variability between samples from the same patient using the shedding profile and a narrower distribution that accounts for residual idiosyncratic noise.

### 2.4. Posterior predictive model assessment

We assess the goodness-of-fit of the models to the data using posterior predictive replication of various summary statistics, the marginal Wanatabe-Akaike information criterion (WAIC) (Merkle et al., 2019), and the ability of the fitted models to predict summary statistics of held-out datasets using posterior predictive validation.

Posterior predictive replication is a useful tool for assessing the fit of a model to data (Gelman et al., 2014a, ch. 6). In short, synthetic replicates of the data generated by sampling from the posterior predictive distribution should not be easily distinguishable from the data the model was fit to. For example, 104 of 148 samples were positive in the composite dataset used to fit the *constant standard* model. If posterior predictive replicates of the data have a similar number of positive samples, the model is able to describe this particular aspect of the data. In contrast, the model would evidently not be suitable if it confidently predicted that all samples should be positive. We assess the goodness-of-fit to the data via posterior predictive replication of two key summary statistics.

The first summary statistics we consider are the number of positive samples *m*_(+)_ and positive patients *n*_(+)_ (i.e. patients with at least one positive sample) because these statistics have been extensively discussed in meta-analyses (Cheung et al., 2020; van Doorn et al., 2020; Wong et al., 2020). As shown in fig. 5 (a) to (d), replicates from all models are consistent with the observed number of positive patients *n*_(+)_ and number of positive samples *m*_(+)_, corroborating our result that the limit of quantification of assays is sufficient to account for negative samples. Replicates from the two *subpopulation* models exhibit larger variability because assay results *x*_*i•*_ for samples from the same patient *i* are correlated due to their mutual dependence on the shedding indicator *z*_*i*_.

**Fig. 5.**
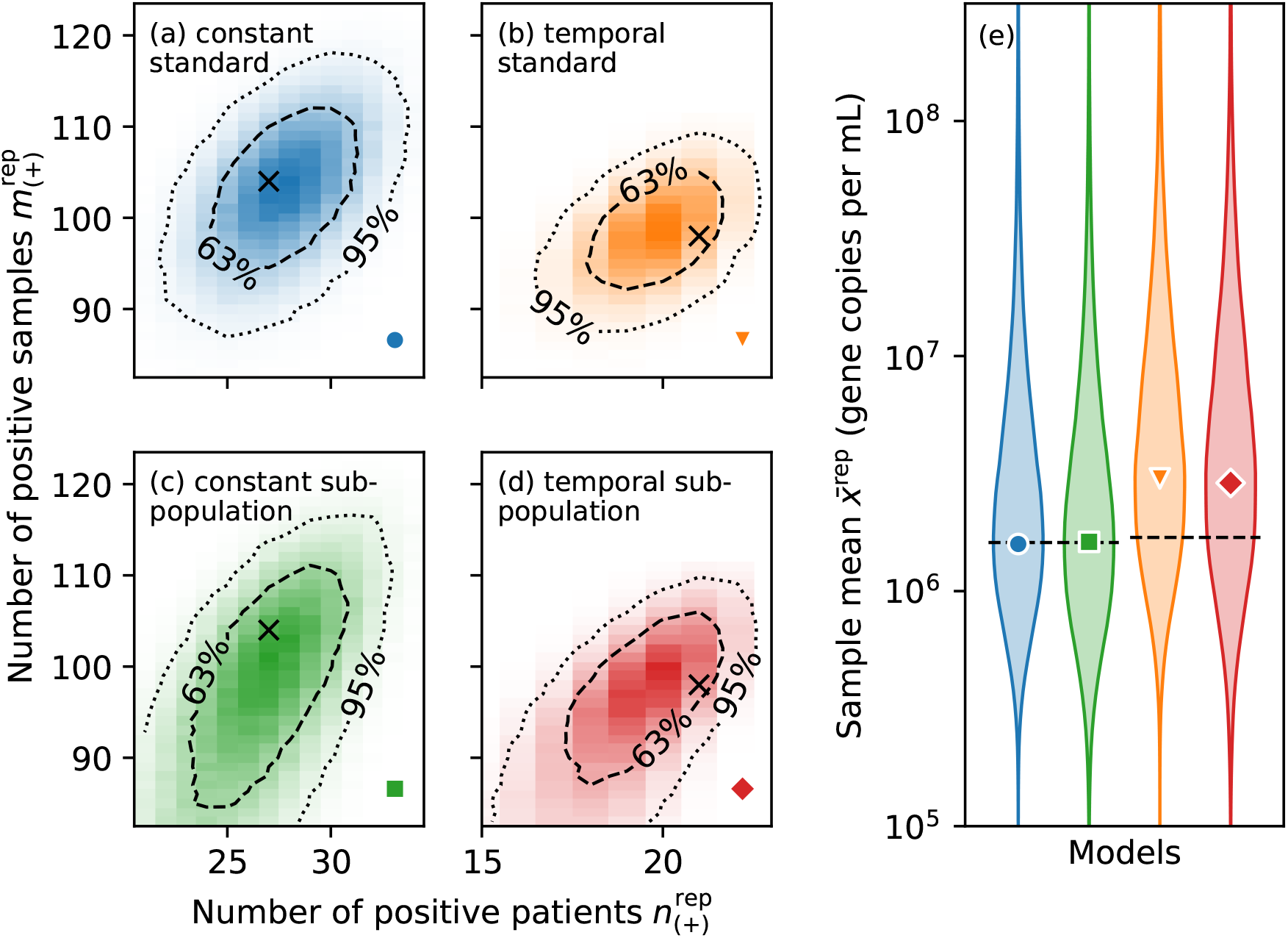
Sampling from the posterior predictive distribution can replicate summary statistics of the data well. Heat maps in panels (a) to (d) represent the joint distribution of the number of positive patients 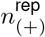 and samples 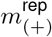 under posterior predictive replication. The observed numbers (shown as black crosses) are consistent with the 63% credible region of the replications. Note that the values differ between the *temporal* and *constant* models because inference for the latter included an additional dataset that did not provide longitudinal information. Panel (e) show posterior predictive replications of the sample mean (excluding negative samples) for all models as violin plots. Markers represent the mode of the distribution, and the observed values are shown as black dashed lines.

Second, we evaluate the sample mean (i.e. the mean of all positive samples) across posterior predictive replicates and compare it with the sample mean of the data. The models can indeed replicate the statistic well, and the observed sample mean is consistent with the predicted sample means, as shown in fig. 5 (e).

In addition to evaluation at the level of the dataset we also sought to evaluate the predictive ability of the four models at the level of individual patients. However, defining an appropriate evaluation criterion is challenging due to the heterogeneity of the data: the level of quantification varies by almost two orders of magnitude between studies, and the number of samples per patient is highly variable. We used the Watanabe-Akaike information criterion (WAIC) because it approximates leave-one-out cross validation of the log predictive density (Gelman et al., 2014b), though with two modifications: first, we employ the marginal WAIC because it is applicable to clustered data, such as samples from the same patient with shared latent mean *λ* (Merkle et al., 2019). Second, we make an adjustment to account for the variable

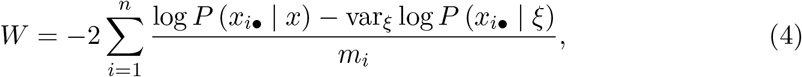

where *P* (*x*_*i•*_ | *x*) is the posterior predictive distribution and var_*ξ*_ denotes the posterior variance with respect to all parameters except the patient mean *λ* which has been marginalised (Merkle et al., 2019). We implemented marginalisation with respect to *λ* using numerical quadrature. To ensure that contributions from patients are on the same scale irrespective of the number of samples *m*_*i*_, we divide by *m*_*i*_ in eq. (4).

Results for the four models are shown in table 2, and standard errors were obtained by bootstrapping. Akin to the log marginal likelihood in section 2.3, the adjusted WAIC strongly prefers *temporal* models. However, it weakly favours models with a *subpopulation* of patients who do not shed RNA in their faeces because these models incur a small penalty ≈ log (1 − *ρ*) for patients without positive samples–even when the shedding prevalence *ρ* is large (see eq. (3)).

Posterior predictive replication and WAIC can only assess the models’ ability to explain the data they were fit to. In other words, a model that satisfies such posterior predictive checks may nevertheless fail to generalise to other datasets (although WAIC includes a correction to account for in-sample evaluation). To assess the out-of-sample predictive utility of the models, we consider predictions for two held-out datasets.

Kim et al. (2020a) collected 129 samples from 38 hospitalised patients, and we assumed that at least one sample was collected from each patient. The remaining 91 samples were assumed to be collected from patients with equal probability. This information is sufficient to generate model-based predictions in two steps by sampling from the posterior predictive distribution. First, we sampled the patient-level means *λ* from the population-level distribution. Second, we sampled the assay results according to eqs. (2) and (3) for the *standard* and *subpopulation* models, respectively. While Kim et al. (2020a) also report the number of positive and negative samples, as shown in table 1, we could not replicate these summary statistics because they do not provide the limit of detection of their assay. Ng et al. (2020) collected 81 samples from 21 patients, and we used the same allocation of samples to patients to generate predictions as for the study by Kim et al. (2020a).

## 3. Results

We fitted each of the four models described in section 2 model to longitudinal data extracted from three studies (Han et al., 2020; Lui et al., 2020; Wölfel et al., 2020) listed in table 1. All studies used RT-qPCR assays to quantify SARS-CoV-2 RNA copies in faecal samples collected from hospitalised patients. The two *constant* models were also fitted to an additional dataset collected by Wang et al. (2020) that does not provide temporal information. As shown in table 2, we compared the models using the Bayesian model evidence (i.e. the marginal likelihood of the data under each model) (Gelman et al., 2014b) and adjusted marginal WAIC, allowing us to draw two conclusions.

First, accounting for the time dependence of the shedding profile is essential. The three shedding profiles we considered are indistinguishable where data are available to constrain them (see section 2.2.1 for details). However, there are only few samples obtained prior to day six past symptom onset, as shown in fig. 2 (a), and shedding behaviour during the early infection course cannot be constrained given the available data. For simplicity, we use the exponential decay profile unless otherwise specified. Typical faecal RNA concentrations decay with a maximum a posteriori half-life of 34 hours (28–43 hours 95% credible interval) among hospitalised patients, as shown in fig. 2 (b) and (c).

Patients may not recall the number of days since symptom onset accurately, or they may present with atypical symptoms that are not easily identified as the onset of COVID19 (Gan et al., 2020). To assess the sensitivity of the inferred half-life to inaccurate reports, we repeated the inference after adding up to three days of reporting noise to the number of days since symptom onset. No sensitivity of the half-life inference to inaccurate reports was observed.

The second result is that there is no evidence for a subpopulation of patients who do not shed viral RNA faecally; *standard* models (without a non-shedding subpopulation) are preferred for both the *constant* (log odds of the marginal likelihood 3.1 ± 0.3) and *temporal* models (log odds of the marginal likelihood 1.8 ± 0.3). Reported errors are standard errors. Consistent with the model comparison results, the inferred shedding prevalence is large and the 95% credible interval includes *ρ* = 1 for both *subpopulation* models. While the adjusted Watanabe-Akaike information criterion marginally favours *subpopulation* models, together, our analysis suggests that the level of quantification of the assays used in the three studies can explain the number of negative patients and samples because “the level of viral RNA present in stool can fluctuate around the margin of laboratory detection” (Ng et al., 2020, p. 642).

To assess the out-of-sample predictive utility of the models, we considered predictions for two held-out datasets that do not provide microdata, as listed in table 1. The studies conducted by Kim et al. (2020a) and Ng et al. (2020) provide sufficiently detailed descriptions of their protocols to simulate the studies and make predictions by sampling from the posterior predictive distribution (see section 2.4 for details). Since our models have not been fit to these data, their ability to predict summary statistics of those data is an indication of how well the models generalise. *Temporal* models are preferred by the data, but we do not have any temporal information about the studies conducted by Kim et al. (2020a) and Ng et al. (2020). Nonetheless, the *constant* models can make accurate out-of-sample predictions because all studies consider the same population: hospitalized patients.

Kim et al. (2020a) collected 129 samples from 38 hospitalised patients, and they reported the largest observed concentration max *x* = 2.7 × 10^7^ gene copies per mL of faeces. Ng et al. (2020) collected 81 samples from 21 patients, and the largest observed RNA concentration was max *x* = 1.3 × 10^7^ copies per mL. As shown in fig. 6 (a) and (b), predictions from our models are consistent with the reported values. Predictions of the maximum are smaller for the study by Ng et al. (2020) than Kim et al. (2020a), which is expected owing to a smaller number of samples (so the tails of the distribution are less well sampled). Ng et al. (2020) also reported the median number of positive samples per patient median *m*_*•*(+)_. Because they report the limit of detection of their assay, we can make predictions about the median number of positive samples per patient which agree with the reported value, as shown in panel (c) of fig. 6.

**Fig. 6.**
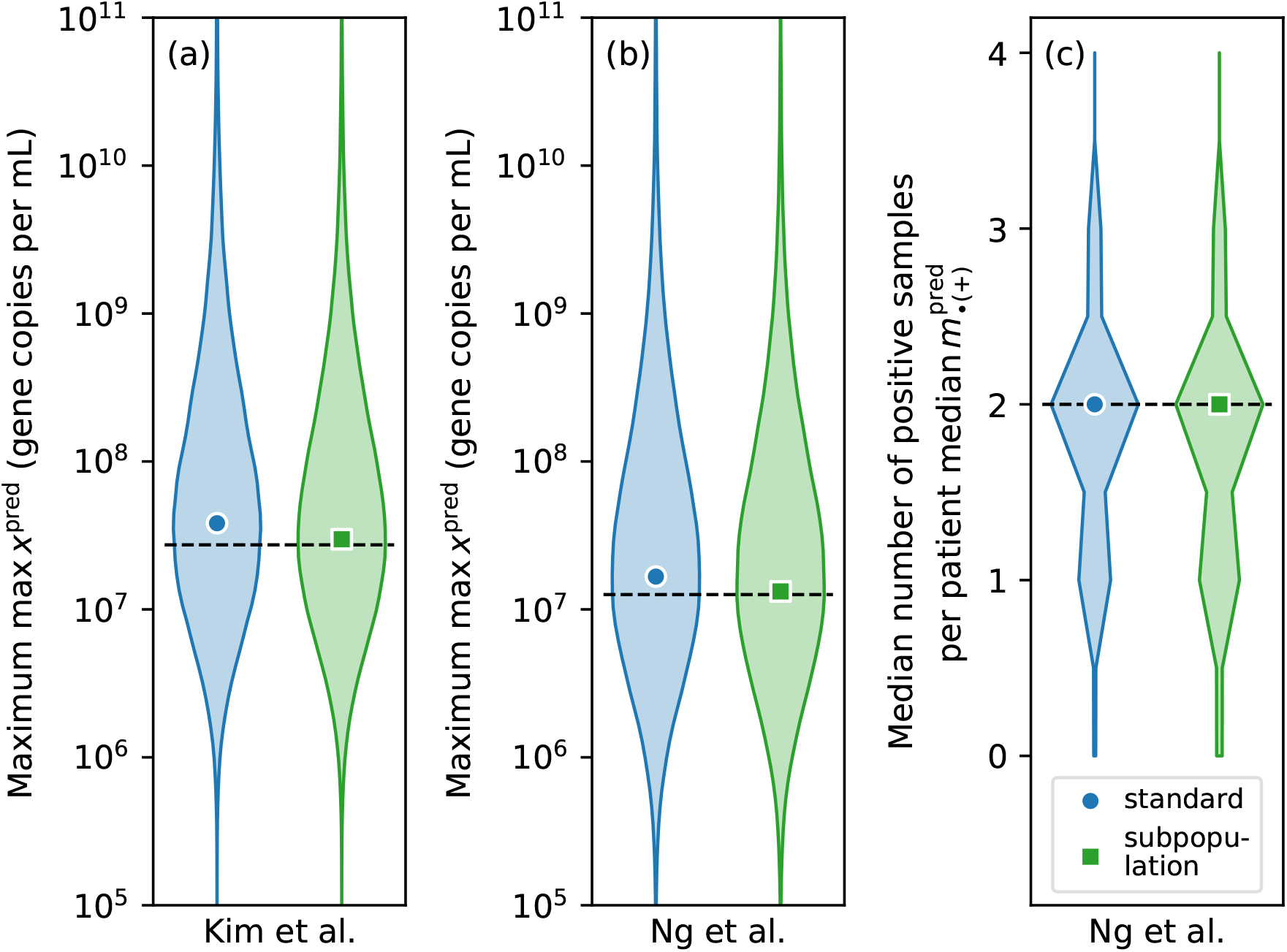
The models can make accurate out-of-sample predictions. Predictions of the maximum concentration observed by Kim et al. (2020a) and Ng et al. (2020) are shown in panels (a) and (b) as violin plots, respectively. The value reported in the studies is shown as black dashed lines. Panel (c) shows predictions of the median number of positive samples per patient reported by Ng et al. (2020). Only results for the *constant* models are shown because the two studies did not provide temporal information.

## 4. Discussion

We have inferred properties of the faecal SARS-CoV-2 RNA shedding distribution by fitting a suite of Bayesian hierarchical models to clinical data from four studies. The models account for the limits of quantification of RT-qPCR assays and a variable number of samples per patient. They are able to capture salient properties of the data and generalise well to two held-out datasets. There is no evidence of patients who do not shed viral RNA faecally.

The inferred temporal shedding profile is robust to inaccurate reports of the number of days since symptom onset, and it suggests that hospitalised patients are in the tail of the shedding profile because faecal RNA concentrations decay by an order of magnitude over the course of four to five days. While extrapolation should be treated with caution, wastewater-based surveillance of COVID-19 lends additional credibility to the hypothesis that SARS-CoV-2 RNA concentrations in wastewater are higher than expected based on faecal shedding inferred from hospitalised patients (Wu et al., 2020). Assuming mean faecal RNA concentrations Λ are not larger in mild cases in the community than among hospitalised patients, a daily per capita wastewater volume of *V* = 300 L (Tscharke et al., 2019), and faecal mass of *m* = 128 g per person per day (Rose et al., 2015), we would expect wastewater RNA concentrations on the order of *m*Λ*/V* ∼ 10^3^ mL^*−*1^ *if every person was infected*. In practice, concentrations in excess of 10^3^ mL^*−*1^ have been observed (Medema et al., 2020) at times when seroprevalence of SARS-CoV-2 antibodies was less than ten percent (Slot et al., 2020). Substantial shedding during the early stages of the infection likely explains these observations. The rapid decay of the shedding profile also implies that signals from wastewater-based surveillance of SARS-CoV-2 are likely more indicative of incidence, rather than prevalence. Wastewater-based surveillance is thus a promising approach for early detection of cases in the community. A better understanding of the shape of the shedding profile, especially prior to symptom onset, is essential for interpreting signals from WBE correctly during critical phases of rapid changes in levels of infection.

While one of the largest quantitative studies revealed no association between disease severity and faecal RNA concentration (Zheng et al., 2020), results obtained from hospitalised patients are unlikely to apply to the general population, e.g. the former tend to be older and have more comorbidities. Faecal samples should be collected from a representative sample of patients over the entire infection course to refine quantitative estimates of faecal shedding of SARS-CoV-2 RNA. These data should include faecal volumes to estimate the total RNA load in faeces in addition to concentrations. The effect of vaccinations and emerging variants on faecal RNA shedding should also be investigated to make wastewater-based surveillance an effective *quantitative* monitoring tool.

## Data Availability

The data analysed during the current study and custom computer code are available at https://github.com/tillahoffmann/shedding.

## Data and code availability

All data and code needed to reproduce the results in this paper are available at https://github.com/tillahoffmann/shedding.

## Acknowledgements

TH was supported by the Natural Environment Research Council under Grant NE/V010387/1. We thank Will Handley for discussions about the *polychord* sampler, Edgar Merkle for feedback on adjusting the marginal WAIC, as well as Nick Jones, Kathleen O’Reilly, Vincent Savolainen, Emma Ransome, Mary Burkitt-Gray, and Christopher Coleman for comments on the manuscript.

## Appendix

We use the parametrisation of the generalised gamma distribution presented by Prentice (1974), and the random variable *x* follows a generalised gamma distribution if

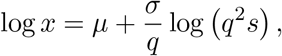

where *s* is a gamma random variable with shape *q*^*−*2^. The probability density function in eq. (2) is thus

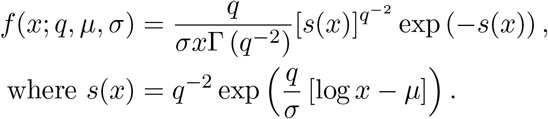

Similarly, the cumulative distribution function is

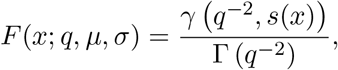

where *γ* is the lower incomplete gamma function.

The expected value is

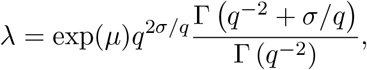

and expressing the location parameter *μ* in terms of the mean *λ* yields eq. (1). The coefficient of variation is

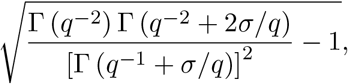

and the generalised gamma distribution has a constant coefficient of variation independent of the location parameter *μ*.

## Notes

### Competing Interest Statement

TH consults the Department for Health and Social Care on statistical analysis of data for wastewater-based surveillance of COVID-19.

### Funding Statement

This work was supported by the Natural Environment Research Council grant number NE/V010387/1.

### Author Declarations

The study was reviewed by the Research Governance and Integrity Team at Imperial College London. Because only publicly available data were used, the study does not require review by the Imperial College Research Ethics Committee.

### Summary of Updates

Update after reviewer comments.

